# S glycoprotein diversity of the Omicron variant

**DOI:** 10.1101/2021.12.04.21267284

**Authors:** Rakesh Sarkar, Mahadeb Lo, Ritubrita Saha, Shanta Dutta, Mamta Chawla-Sarkar

## Abstract

On the backdrop of ongoing Delta variant infection and vaccine-induced immunity, the emergence of the new Variant of Concern, the Omicron, has again fuelled the fears of COVID-19 around the world. Currently, very little information is available about the S glycoprotein mutations, transmissibility, severity, and immune evasion behaviour of the Omicron variant. In the present study, we have performed a comprehensive analysis of the S glycoprotein mutations of 309 strains of the Omicron variant and also discussed the probable effects of observed mutations on several aspects of virus biology based on known available knowledge of mutational effects on S glycoprotein structure, function, and immune evasion characteristics.

## Introduction

Increased transmissibility and high mutation rate of SARS-CoV-2, the causative agent of COVID-19, led to the emergence of multiple variants of concern (VOCs) characterized by the presence of genetic changes which are known to affect virus characteristics such as transmissibility, disease severity, immune escape, and diagnostic or therapeutic escape. Till mid-November 2021, there were four VOCs called Alpha (B.1.1.7), Beta (B.1.351), Gamma (P.1), and Delta (B.1.617.2). On 26 November 2021, WHO designated the SARS-CoV-2 variant B.1.1.529 as a new VOC and named it Omicron. This decision was made based on the advice of WHO’s Technical Advisory Group on Virus Evolution (TAG-VE) recommendation as the Omicron has numerous mutations, some of which are shared with other VOCs, that may enhance transmissibility, disease severity, immune escape, and diagnostic or therapeutic escape, though no direct evidence exists. WHO was informed about the emergence of the variant B.1.1.529 on 24 November, 2021 from South Africa. South Africa is currently dealing with the third wave of COVID-19, which is mainly dominated by the Delta variant. Infections have increased dramatically in recent weeks in South Africa, coinciding with the discovery of the B.1.1.529 strain. However, whether this increased case of COVID-19 is due to the Omicron or other factors needs to be verified by genome sequencing and epidemiological studies. The first known infection of B.1.1.529 was confirmed from a specimen collected on 9 November 2021 [**1, 2**].

SARS-CoV-2 genome codes for multiple proteins, including the spike (S) glycoprotein that protrudes from the virus envelope [**3**]. The S glycoprotein plays crucial role in the very early stage of virus life cycle that includes virus attachment to the host cell surface, membrane fusion and entry into the host cell [**3-10**]. The S glycoprotein, as a surface protein, is the primary target of neutralizing antibodies elicited by the host adaptive immune response [**4, 11-14**]. In the constant tug of war between the host and the virus, virus strains with S glycoprotein mutations that facilitate virus entry and/or help the virus escape neutralizing antibodies are frequently selected and eventually predominate. In the lights of crucial role of S glycoprotein in virus infection and host immune evasion, scientists have prioritized mutations that have been emerged within the S glycoprotein of circulating SARS-CoV-2 strains and also investigated the biological significance of those mutations [**15, 16**]. In the present study, we performed a comprehensive analysis of S glycoprotein mutations of the Omicron variant and also classified them into different groups based on different combination of coexisting S glycoprotein mutations.

## Materials and Methods

### Retrieval of genome sequences of the Omicron variant deposited in GISAID

For retrieval of SARS-CoV-2 genome sequences of the Omicron variant, we accessed to Global Initiative on Sharing All Influenza Data (GISAID) on 2 December 2021 [**17**]. By applying filter on Variant (VOC Omicron GR/484A), we observed that a total 309 genome sequences of the Omicron variant were submitted. We downloaded all these genome sequences from GISAID for further analysis. Name of all the Omicron variants are presented in **Table S1**. The genome sequences of the prototype SARS-CoV-2 strain hCoV-19/Wuhan/WIV04/2019 (GISAID accession no. EPI_ISL_402124) was also downloaded from the GISAID database for the purpose of mutational analysis.

### Mutational analysis of the S glycoprotein

For performing mutational analysis, the S glycoprotein protein coding region of 309 genome sequences of the Omicron variant as well as the prototype genome (hCoV-19/Wuhan/WIV04/2019) were translated to amino acid sequences by using TRANSEQ nucleotide-to-protein sequence conversion tool (EMBL-EBI, Cambridgeshire, UK). Next, the S glycoprotein protein sequences of all the SARS-CoV-2 strains including the prototype variant and the Omicron variantswere aligned by MEGA software (Version X) and subsequently observed for amino acid substitutions in the S protein of the Omicron variants with compared to the prototype strain [**18**]. The amino acid substitution observed at a particular location of the S glycoprotein of the Omicron variant was marked with the number according to its position with reference to the first amino acid of the S glycoprotein of the prototype strain.

### Phylogenetic analysis

The phylogenetic analysis of the 155 sequences of the Omicron variant was performed with the Ultafast Sample Placement of Existing Trees (UShER) that has been integrated in the UCSC SARS-CoV-2 Genome Browser [**19**]. We accessed to the UCSC SARS-CoV-2 Genome Browser (https://genome.ucsc.edu/cgi-bin/hgPhyloPlace) and uploaded the name of 155 strains (**Table S2**) for the construction of the phylogenetic tree. UShER is a program that rapidly places new samples onto an existing phylogeny using maximum parsimony. It is particularly helpful in understanding the relationships of newly sequenced SARS-CoV-2 genomes with each other and with previously sequenced genomes in a global phylogeny.

## Results

### Geographical distribution of the Omicron variant

Geographical distribution showed that 309 sequences of the Omicron variant have been deposited to the GISAID from 21 different nations across six continents (**Table 1**). Majority of these sequences (N=223) were submitted from Africa, which includes 4 representing countries South Africa (N=170), Ghana (N=33), Botswane (N=19) and Reunion (N=1). There were 62 sequences submitted from 11 different countries (United Kingdom, N=18; Portugal, N=13; Netherlands, N=12; Austria, N=5; Germany, N=5; Italy, N=3; Belgium, N=1; Czech Republic, N=1; Spain, N=1; Sweden, N=1; Ireland, N=1) in Europe. Among the rest 24 sequences, 9 were submitted from three Asian countries (Hong Kong, N=6; Japan, N=2; Israel, N=1), 9 from Australia of Oceania, 3 from Canada of North America, and 3 from Brazil of South America.

**Table 1:** Geographical distribution of the Omicron variant

| Continents | Country | Frequency | Total frequency |
| --- | --- | --- | --- |
| Africa | South Africa | 170 | 223 |
|  | Ghana | 33 |  |
|  | Botswana | 19 |  |
|  | Reunion | 1 |  |
| Europe | United Kingdom | 18 | 62 |
|  | Portugal | 13 |  |
|  | Netherlands | 12 |  |
|  | Austria | 5 |  |
|  | Germany | 5 |  |
|  | Italy | 3 |  |
|  | Belgium | 2 |  |
|  | Czech Republic | 1 |  |
|  | Spain | 1 |  |
|  | Sweden | 1 |  |
|  | Ireland | 1 |  |
| Asia | Hong Kong | 6 | 9 |
|  | Japan | 2 |  |
|  | Israel | 1 |  |
| Oceania | Australia | 9 | 9 |
| North America | Canada | 3 | 3 |
| South America | Brazil | 3 | 3 |
| Worldwide | 21 different countries |  | 309 |

### Mapping of the S glycoprotein mutations of the Omicron variant

Mutational analysis of the S glycoprotein of the Omicron variant revealed the presence of 37 dominant mutations which ranges in the frequency from 59% to 100% (**Table 2**). The S1 domain of the S glycoprotein contains 31 different mutations that encompasses 11 mutations (A67V, ΔH69, ΔV70, T95I, G142D, ΔV143, ΔY144, ΔY145, ΔN211, L212I and ins214EPE) within the N-terminal domain (NTD), 15 mutations (G339D, S371L, S373P, S375F, K417N, N440K, G446S, S477N, T478K, E484A, Q493R, G496S, Q498R, N501Y and Y505H) within the Receptor binding domain (RBD) and 5 mutations (T547K, D614G, H655Y, N679K and P681H) at the C-terminus of the S1 subunit. Interestingly, among the 15 mutations of the RBD, 10 mutations (N440K, G446S, S477N, T478K, E484A, Q493R, G496S, Q498R, N501Y and Y505H) were observed within the Receptor binding motif (RBM). The S2 subunit of the S glycoprotein was found to have 6 mutations, of which N764K and D796Y were present at the N-terminus of the S2 subunit, N856K was found within the region between Fusion peptide (FP) and Heptad repeat sequence 1 (HR1), and Q954H, N969K, and L981F were present within the HR1 (**Figure 1**). By comparing the S glycoprotein mutations of four VOCs Alpha, Beta, Gamma, and Delta, it has been observed that the Omicron contains 25 unique mutations (A67V, ΔV143, ΔN211, L212I, ins214EPE, G339D, S371L, S373P, S375F, N440K, G446S, S477N, E484A, Q493R, G496S, Q498R, Y505H, T547K, N679K, N764K, D796Y, N856K, Q954H, N969K, and L981F), whereas 12 mutations (ΔH69, ΔV70, T95I, G142D,ΔY144, ΔY145, K417N, T478K,N501Y, D614G, H655Y, and P681H) were shared with other four VOCs (**Figure 1**).

**Table 2:** Frequency of different mutations associated with the S glycoprotein of the Omicron variant (B.1.1.529)

| Number | Mutations | Frequency (N=309) | Percentage (%) |
| --- | --- | --- | --- |
| 1 | A67V | 306 | 99.03 |
| 2 | ΔH69 | 296 | 95.79 |
| 3 | ΔV70 | 294 | 95.15 |
| 4 | T95I | 304 | 98.38 |
| 5 | G142D | 285 | 92.23 |
| 6 | ΔV143 | 286 | 92.56 |
| 7 | ΔY144 | 285 | 92.23 |
| 8 | ΔY145 | 287 | 92.88 |
| 9 | ΔN211 | 262 | 84.79 |
| 10 | L212I | 262 | 84.79 |
| 11 | Ins214EPE | 258 | 83.50 |
| 12 | G339D | 304 | 98.38 |
| 13 | S371L | 274 | 88.67 |
| 14 | S373P | 274 | 88.67 |
| 15 | S375F | 266 | 86.08 |
| 16 | K417N | 183 | 59.22 |
| 17 | N440K | 199 | 64.40 |
| 18 | G446S | 217 | 70.23 |
| 19 | S477N | 289 | 93.53 |
| 20 | T478K | 289 | 93.53 |
| 21 | E484A | 289 | 93.53 |
| 22 | Q493R | 288 | 93.20 |
| 23 | Q496S | 279 | 90.29 |
| 24 | Q498R | 287 | 92.88 |
| 25 | N501Y | 285 | 92.23 |
| 26 | Y505H | 283 | 91.59 |
| 27 | T547K | 308 | 99.68 |
| 28 | D614G | 309 | 100.00 |
| 29 | H655Y | 308 | 99.68 |
| 30 | N679K | 306 | 99.03 |
| 31 | P681H | 306 | 99.03 |
| 32 | N764K | 232 | 75.08 |
| 33 | D796Y | 303 | 98.06 |
| 34 | N856K | 304 | 98.38 |
| 35 | Q954H | 304 | 98.38 |
| 36 | N969K | 298 | 96.44 |
| 37 | L981F | 296 | 95.79 |

**Figure 1:**
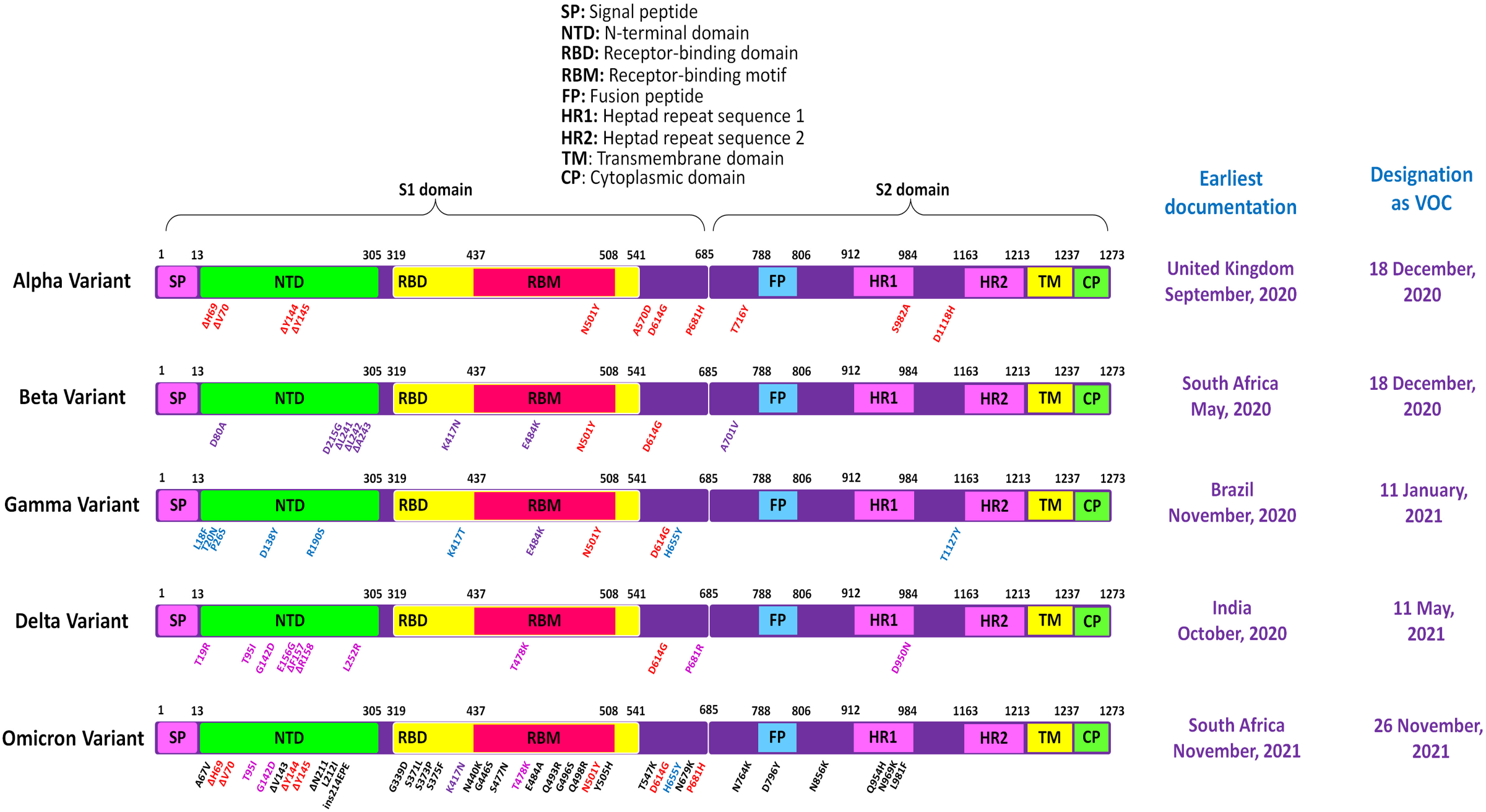
Mapping of the S glycoprotein mutations of the five different VOCs Alpha, Beta, Gamma, Delta and Omicron. The Omicron variant S glycoprotein contains 37 dominant mutations, of which 12 overlap with 4 other VOCs and 25 are novel.

Based on coexisting mutations, we also classified 309 strains of the Omicron variant into 66 different groups, each group representing a different set of coexisting S glycoprotein mutations (**Table 3, Supplementary file 1**). More than half of the Omicron variants (N=170) were found to belong within Group 1, with all the 37 different mutations, and Group 2, with all the mutations except K417N, N440K, G446S and N764K (Group1, N=118; Group 2, N=52). Whereas, rest of the 64 groups represented only 139 strains. Presence of multiple groups demonstrated the high mutational diversity of the S glycoprotein of the Omicron variant.

**Table 3:**
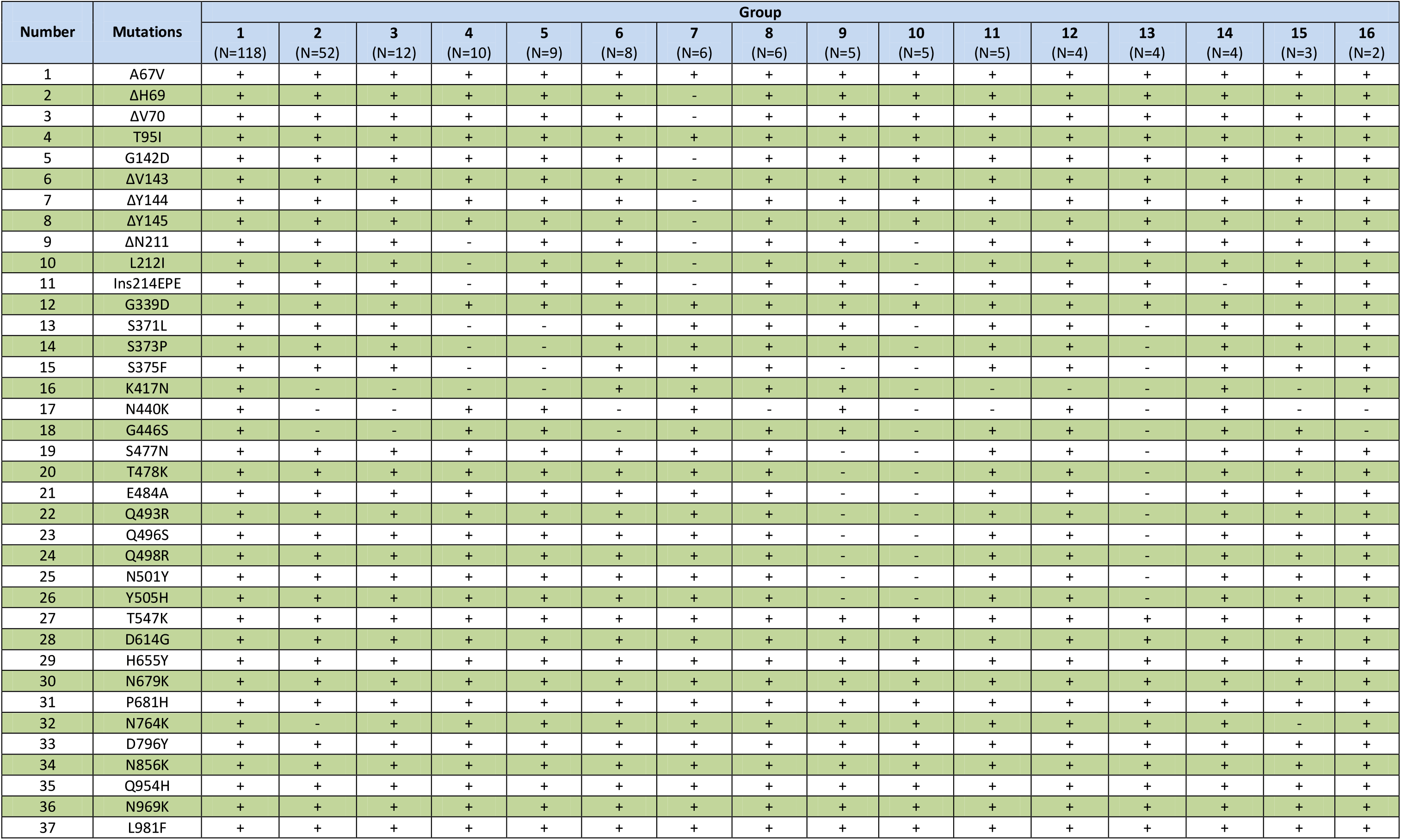
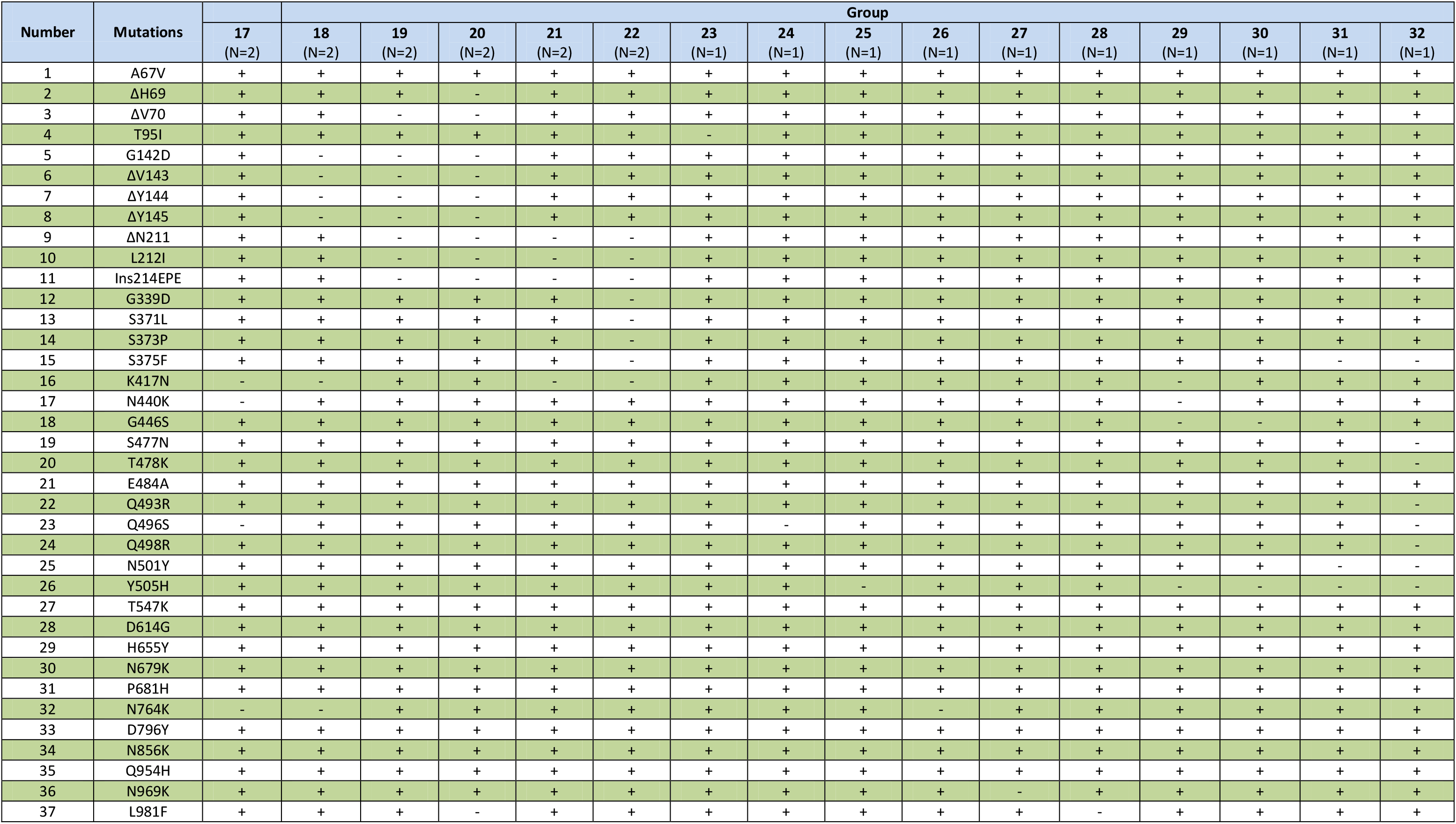
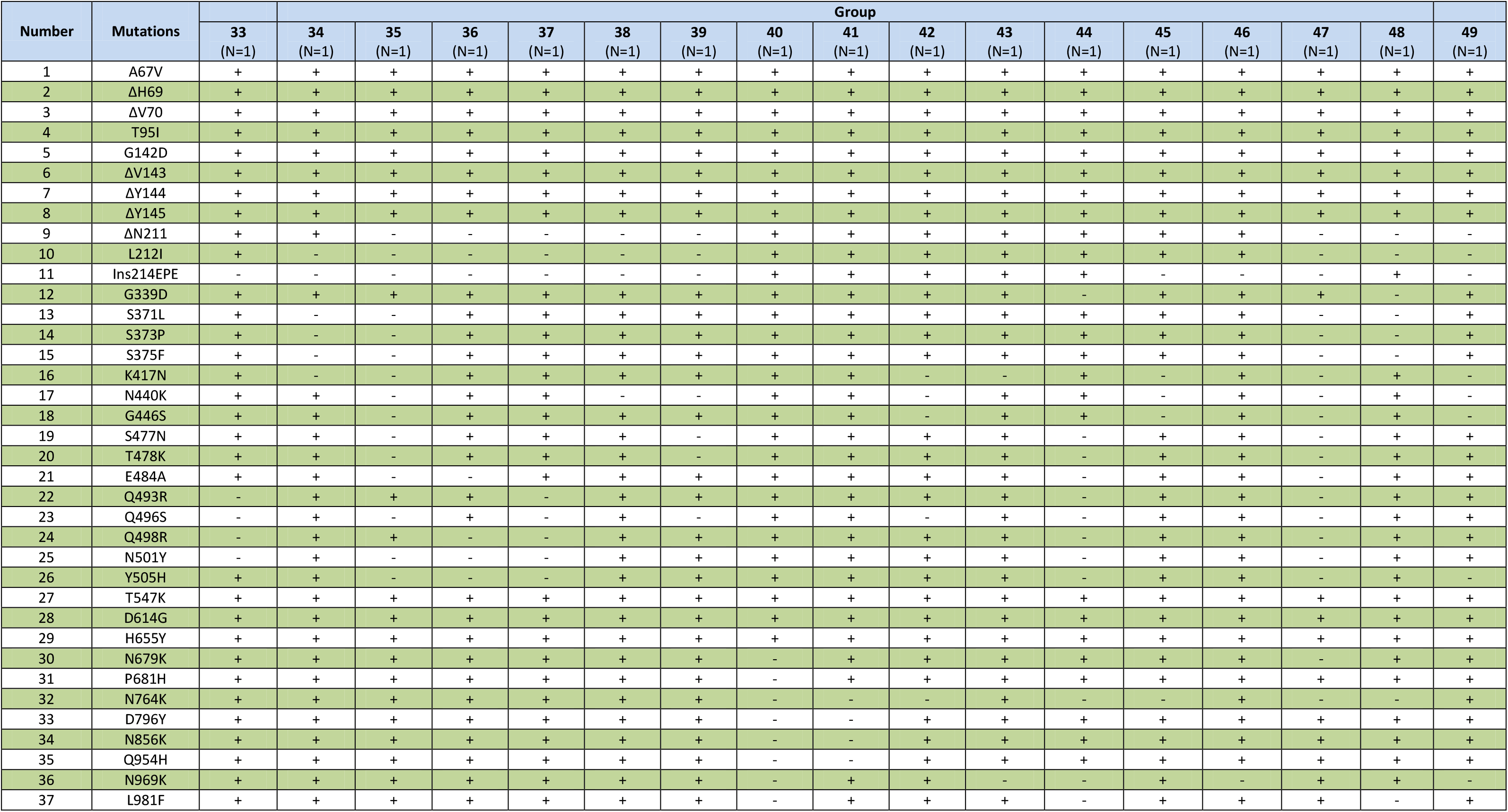
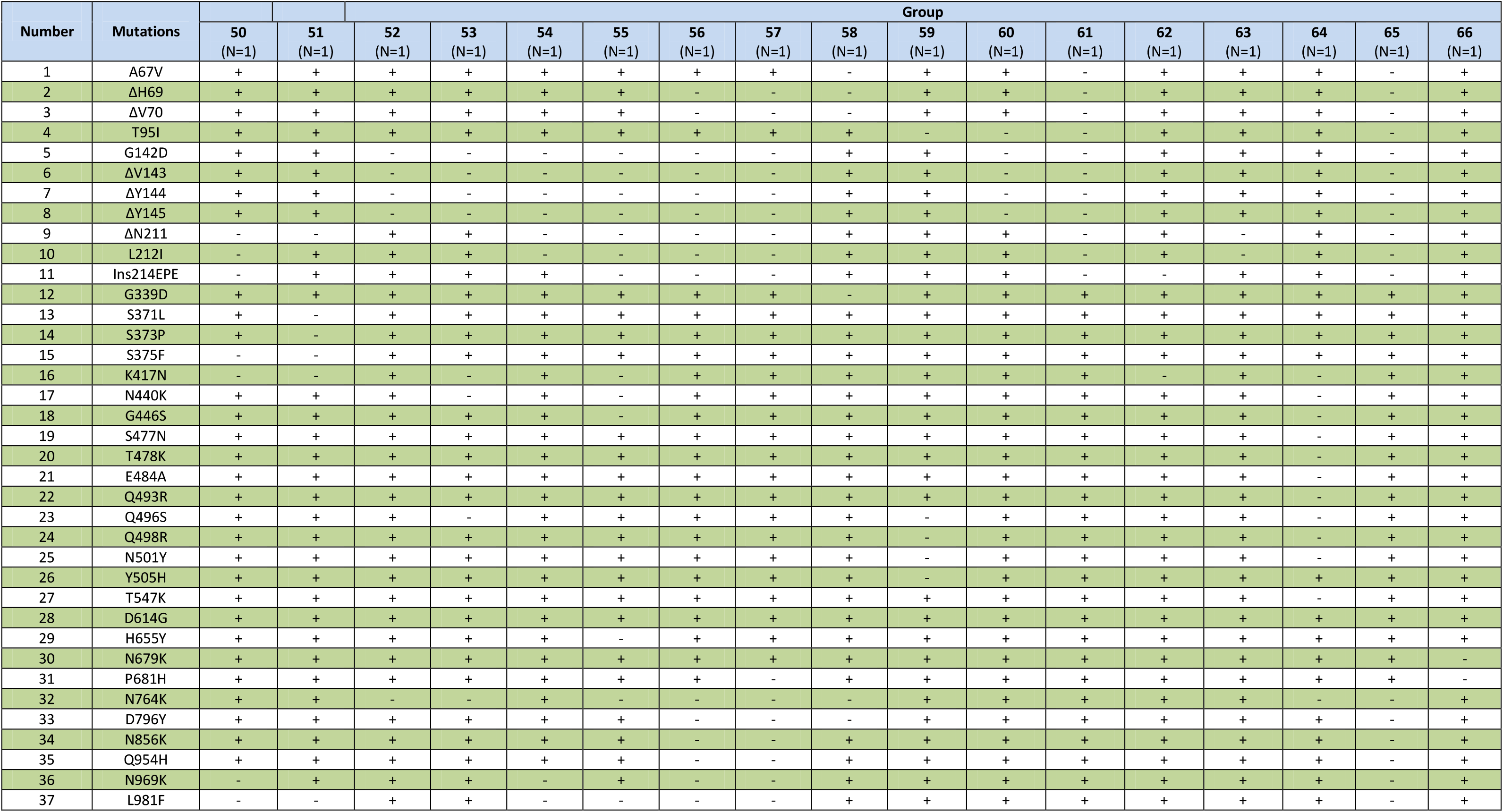
Different groups of Omicron variant having different combination of S glycoprotein mutations

### Phylogenetic analysis of the Omicron variant

We performed phylogenetic analysis of a total 2160 SARS-CoV-2 strains which includes 155 strains of the Omicron variant from 14 different countries and 2005 strains from 19 different clades (20H/Beta, N=13; 20I/Alpha, N=475; 20J/Gamma, N=38; 20A/Delta, N=1049; 21B/Kappa, N=2; 21C/Epsilon, N=29; 21D/Eta, N=6; 21F/Iota, N=19; 21G/Lambda, N=5; 21H, N=5; 19A, N=6; 19B, N=4; 20A, N=97; 20E/EU1, N=65; 20G, N=47; 20C, N=54; 20B, N=77; 20D, N=3; 20F, N=10) by UShER. The metadata of the phylogenetic tree has been provided in the **Supplementary file 2**. The dendrogram showed that 155 strains of the Omicron variant formed a new cluster that emerged from the 20B clade (also known as GR clade or Pango lineage B.1.1). Interestingly, this new cluster of the Omicron variant further divided into multiple sub-clusters depending on the coexisting mutations of the S glycoprotein (**Figure 2)**.

**Figure 2:**
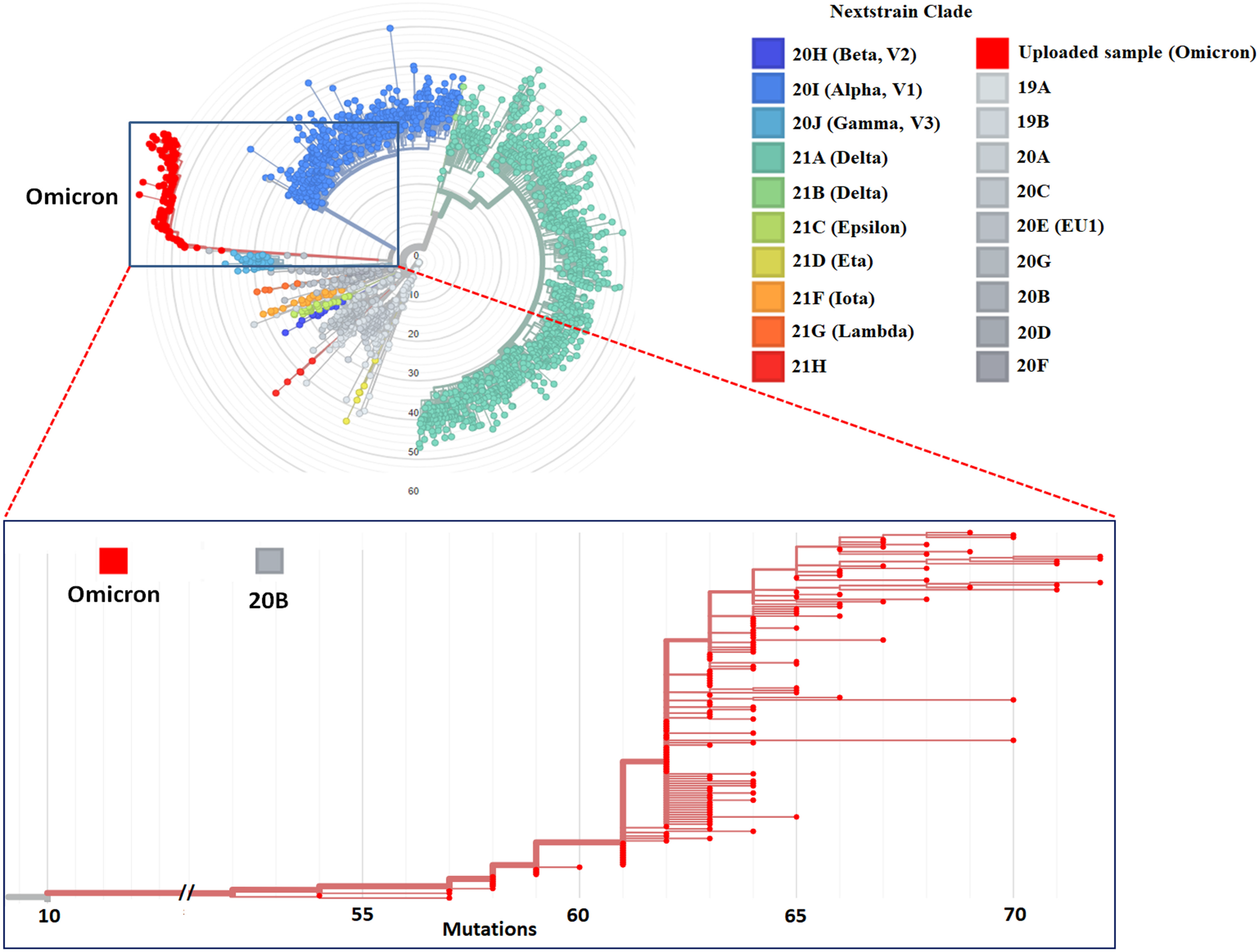
Molecular phylogenetic analysis of new variant strains by UShER. The phylogenetic tree (Radial) was constructed with total 2160 SARS-CoV-2 strains involving 155 strains of the Omicron variant (red color) and 2005 strains from 19 different clades. Each color in tree is representing a different clade/lineage.

## Discussion

Presence of more than 35 mutations in the S glycoprotein, especially in the NTD and RDB of the S1 subunit, of the Omicron variant has again fuelled the fears of COVID-19 around the world. The S glycoprotein, which mediates viral attachment to ACE2 receptor and entry into the host cell, is subdivided into two functional subunits, known as S1 and S2, which form non-covalent interaction after being cleaved by furin during synthesis [**3, 4**]. The RBD and NTD are two crucial domains of the S1 subunit that are responsible for interacting with host cell receptor (ACE2) and recognizing various attachment factors, respectively [**3-9**]. The fusion mechanism is housed in the S2 subunit, which undergoes large-scale conformational changes to force fusion of the virus and host membranes, allowing genome delivery and initiation of infection [**10**]. As RBD domain is immuno-dominantand also required for ACE2 attachment, any mutation in RBD domain could affect the neutralization efficacy of antibodies generated in convalescent and vaccinated individual as well as the binding affinity of the S glycoprotein to ACE2 receptor [**4, 11-14**]. There are 37 dominant mutations in the S glycoprotein of the Omicron variant, raising concerns about whether it is more infectious or pathogenic than other four VOCs, and whether it can evade the natural immunity or vaccine-induced immunity. Despite the lack of definitive immunological and clinical data, we can provide preliminary indications on pathogenicity, transmissibility, and immune evasion capabilities of the Omicron variant based on the known impact of previously identified mutations. Twelve mutations of the Omicron variant (ΔH69, ΔV70, T95I, G142D, ΔY144, ΔY145, K417N, T478K, N501Y, D614G, H655Y, and P681H) overlap with those in the Alpha, Beta, Gamma, and Delta. All these mutations have previously been linked with high transmissibility, increased viral binding affinity, and immune evasion [**15, 20-22**]. Higher transmissibility and increased immune evasion are anticipated from the Omicron variant if these overlapping VOCs mutations maintain their known effects. The functional implications of the remaining 25 mutations (A67V, ΔV143, ΔN211, L212I, ins214EPE, G339D, S371L, S373P, S375F, N440K, G446S, S477N, E484A, Q493R, G496S, Q498R, Y505H, T547K, N679K, N764K, D796Y, N856K, Q954H, N969K, and L981F) of the Omicron variant are unknown, leaving a lot of questions about how the whole set of mutations may affect viral fitness and vulnerability to natural and vaccine-mediated immunity.

Several studies on epitope mapping and antibody foot printing have showed that serum neutralizing antibodies of infected and vaccinated individuals mainly target RBD domain of the S glycoprotein [**11, 23-26]**. Among the 15 mutations of the RBD of the Omicron variant, role of only 5 mutations (K417N, K477N, T478K, E484A and N501Y) have previously been described in the context of immune evasion. The K417N, previously detected in Beta and Delta plus, has been shown to reduce the neutralization efficacy of some monoclonal antibodies [**27, 28**]. Residues E484 and T478 are the part of immuno-dominant site of RBD [**11-13**]. The E484K previously observed in Beta and Gamma, has been shown escape antibody neutralization, and also been found to emerge as escape mutation during exposure to monoclonal antibodies and convalescent plasma [**29-31]**. Four mutant viruses with E484A, E484D, E484G and E484K were also found to be resistant against neutralization by each of the four convalescent sera tested [**32]**. The E484Q has been shown to reduce serum neutralizing antibody titers [**33-35**]. However, the T478K, previously detected in Delta, does not affect neutralization by monoclonal antibodies [**16**]. The K477N has been shown to confer resistant against monoclonal antibodies, but not convalescent plasma [**32**]. The pseudotyped virus carrying N501Y mutation, previously observed in Alpha, Beta and Gamma, also showed resistance to neutralization by monoclonal antibodies [**36**]. Therefore, the presence of four immune escape mutations K417N, K477N, E484A and N501Y in the RBD is likely to improve the immune evasion ability of the Omicron variant. Functional significance of the remaining 10 novel mutations (G339D, S371L, S373P, S375F, N440K, G446S, Q493R, G496S, Q498R, and Y505H) of the RBD in immune evasion needs to be evaluated. Importantly, in all these substitutions, there were significant changes in hydrophobicity of amino acids which may alter the neutralization sensitivity of the virus to monoclonal antibodies or vaccine-induced serum antibodies by altering the epitope structure of the RBD. Although RBD domain is immune-dominant, NTD of the S glycoprotein can also elicit antibody response upon infection and vaccination [**8, 37, 38]**. The NTD domain contains an ‘antigenic supersite’ which comprises of N-terminus (residues 14-20), a β-hairpin (residues 140-158) and a loop (residues 245-264) [**8**]. Among 11 mutations of the NTD of the Omicron variant, 4 mutations (G142D, ΔV143, ΔY144, ΔY145) reside within the β-hairpin region of the antigenic supersite and likely to contribute immune evasion significantly. The role of other 7 mutations (A67V, ΔH69, ΔV70, T95I, Δ ΔN211, L212I and ins214EPE) remains elusive and requires urgent study.

A recent study has illustrated the functional significance of all the RBD mutations of SARS-CoV-2 on ACE2 binding [**39**]. Among the 15 mutations found within the RBD domain of the Omicron variant, 4 mutations (G339D, N440K, T478K and N501Y) were demonstrated to enhance the affinity of RBD towards ACE2, whereas rest 11 mutations (S371L, S373P, S375F, K417N, G446S, S477N, T478K, E484A, Q493R, G496S, Q498R, and Y505H) were demonstrated to reduce the affinity of RBD for ACE2. Notably, mutations at Q493, Q498 and N501 are very crucial for RBD and ACE2 interactions because residues Q493, Q498 and N501 of RBD participate in polar contact networks involving the ACE2 interaction hotspot residues K31 and K353. Amino acid substitutions with nonpolar amino acid at these sites enhance the affinity of RBD to ACE2 [**39**]. However, in the Omicron variant, glutamine (Q) substitution with more polar amino acid arginine (R) at positions 493 and 498 is suspected to reduce the affinity of RBD to ACE2. In contrast, substitution of asparagine at position 501 with less polar amino acid tyrosine will enhance the affinity of RDB to ACE2. The overall affinity of RBD of the Omicron variant to ACE2 will be determined by the magnitude of 4 affinity enhancing mutations and 11 affinity reducing mutations.

Five mutations, including 2 functionally known mutations D614G and P681H, were observed outside the RBD and NTD of the S1 subunit. The Omicron variant is expected to maintain the high transmissibility due to the presence of D614G and P681H mutations, which were previously described key mutations for enhanced transmissibility and infectivity of the virus [**22, 40**]. The S2 subunit of the Omicron variant contains 6 mutations, 3 (Q954H, N969K, and L981F) of which are present within the HR1. Following binding of RBD to the ACE2 receptor on the host cell, HR1 and HR2 domains of the S2 subunit interact to form six-helix-bundle (6-HB) which brings the viral membrane and the host membrane in close proximity, leading to membranes fusion and initiation of infection [41]. Mutations Q954H, N969K, and L981F may enhance the interaction affinity between HR1 and HR2, leading to augmented membrane fusion and infectivity. Currently, it is not clear whether the Omicron has higher transmissibility than the Delta. Although preliminary data suggests that the Omicron variant is spreading rapidly against a backdrop of ongoing Delta variant infection and natural as well as vaccine-induced immunity, indicating high transmissibility and potency to make breakthrough infections. If the current trend continues, the Omicron variant will supplant delta as the most common variant in South Africa and other part of the world very rapidly and may lead to another wave of COVID-19.

## Data Availability

All data produced in the present study are available upon reasonable request to the authors

## Acknowledgement

We would like to acknowledge the scientists and researchers for their valued contribution in SARS-CoV-2 genome sequencing and deposition in GISAID. We would also like to applaud GISAID consortium for allowing us the open access to the deposited SARS-CoV-2 sequences.

## Author contribution

RS and MCS conceived and designed the research. RS, ML and RS performed sequence retrieval, mutational analysis, and figure and table preparation. SD and MCS guided the project and gave valuable scientific inputs. RS and MCS wrote the manuscript. All authors read and approved the manuscript.

---

**Supplementary Table 1:**
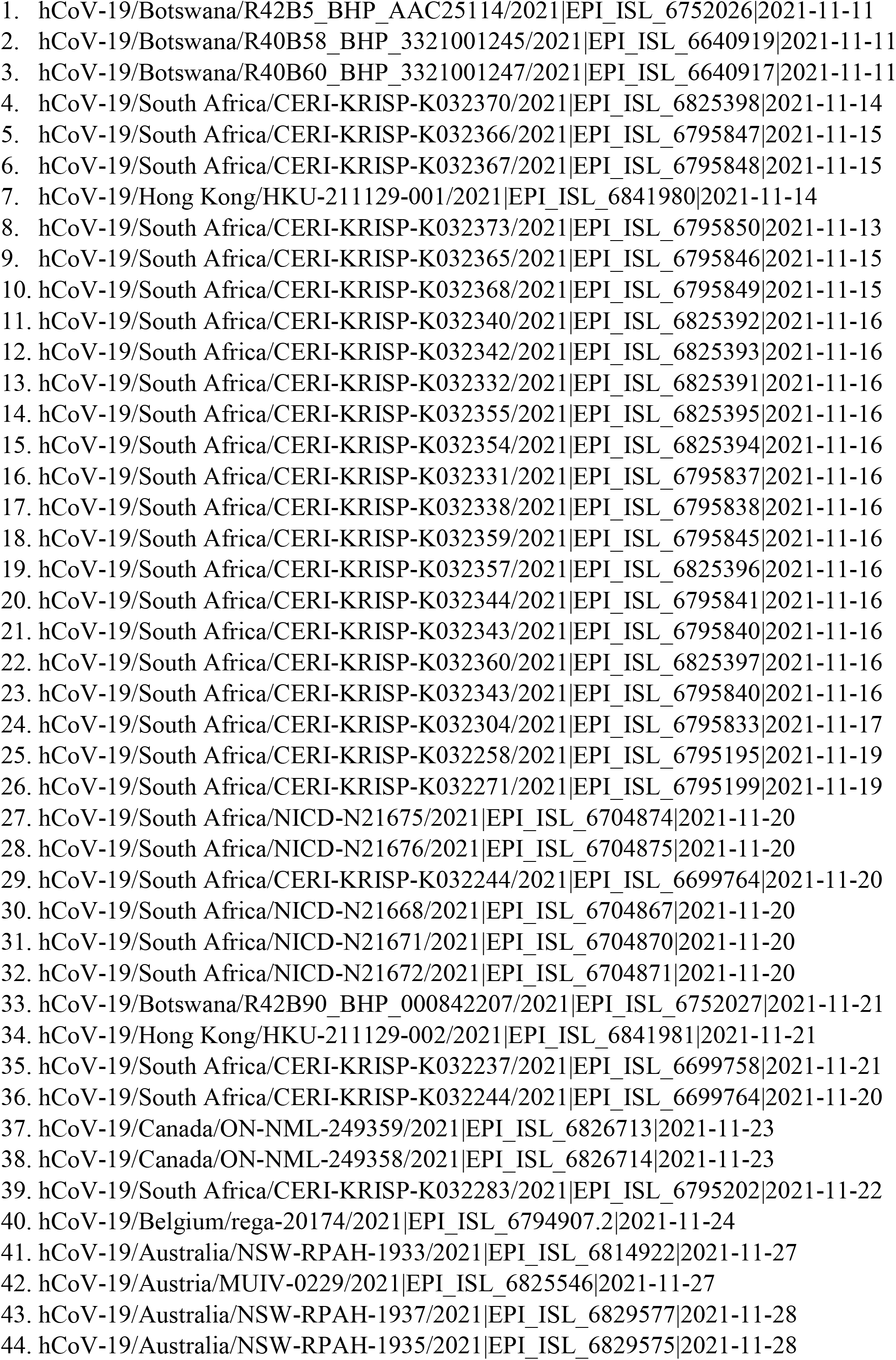
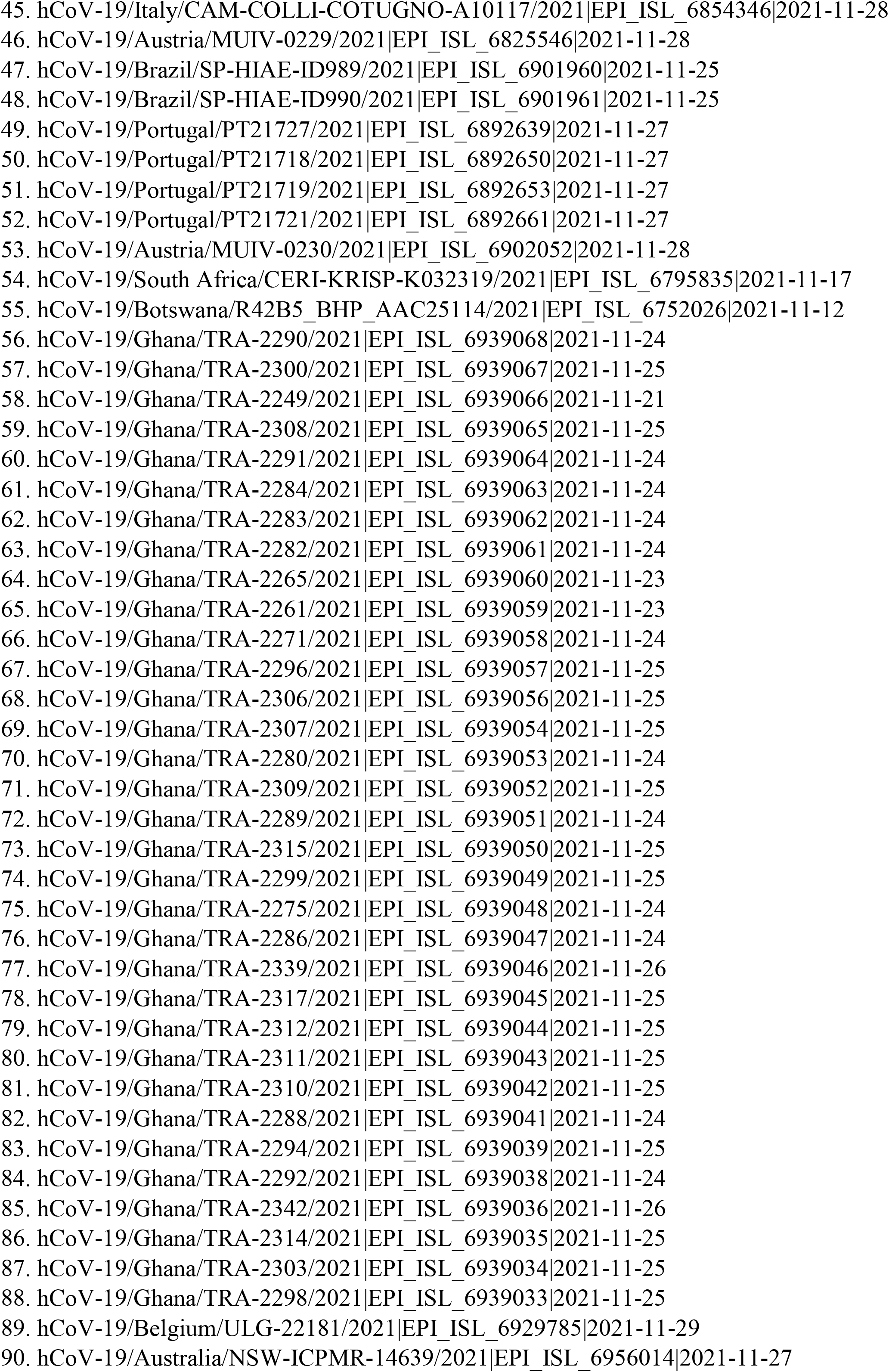
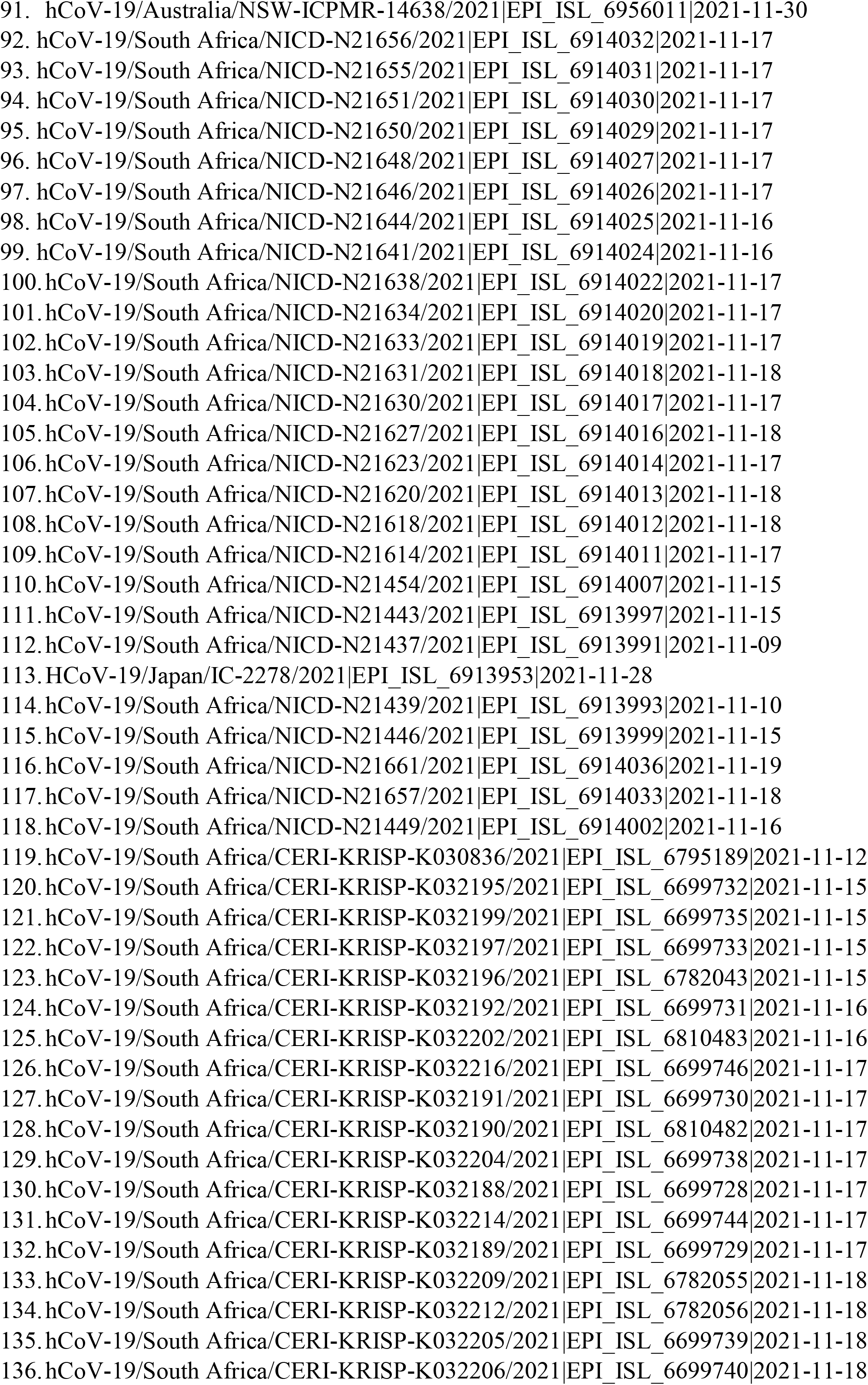
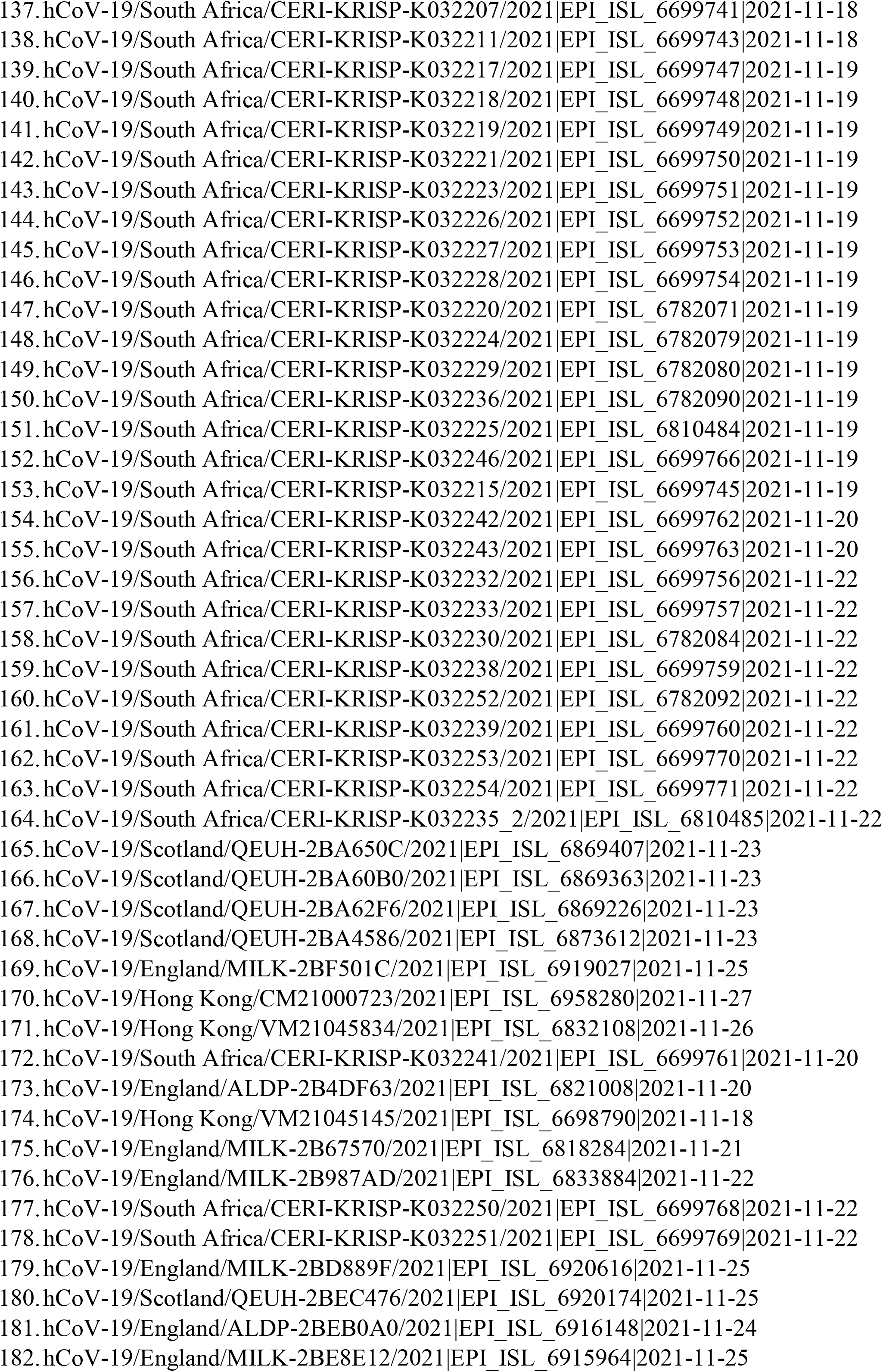
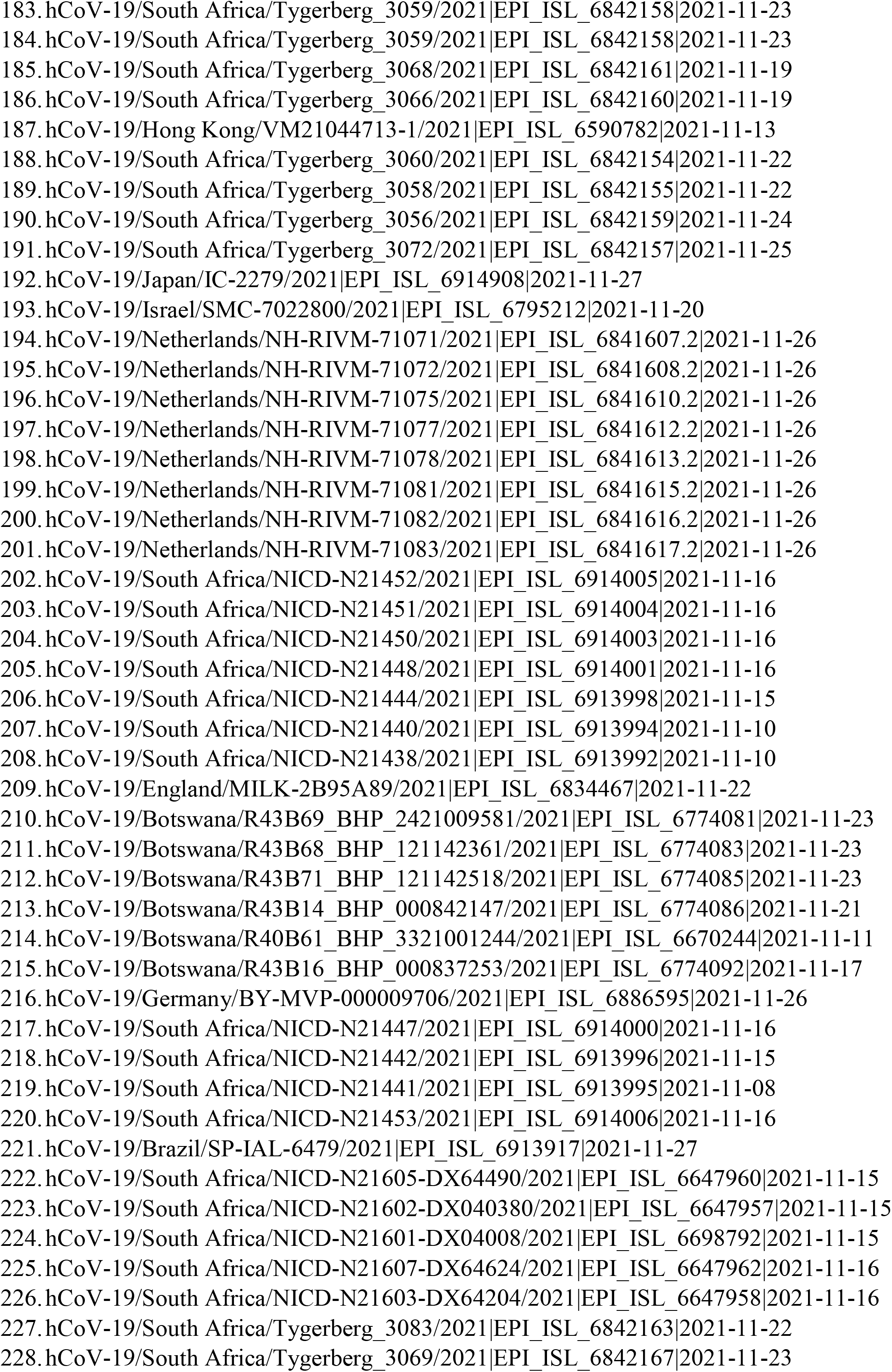
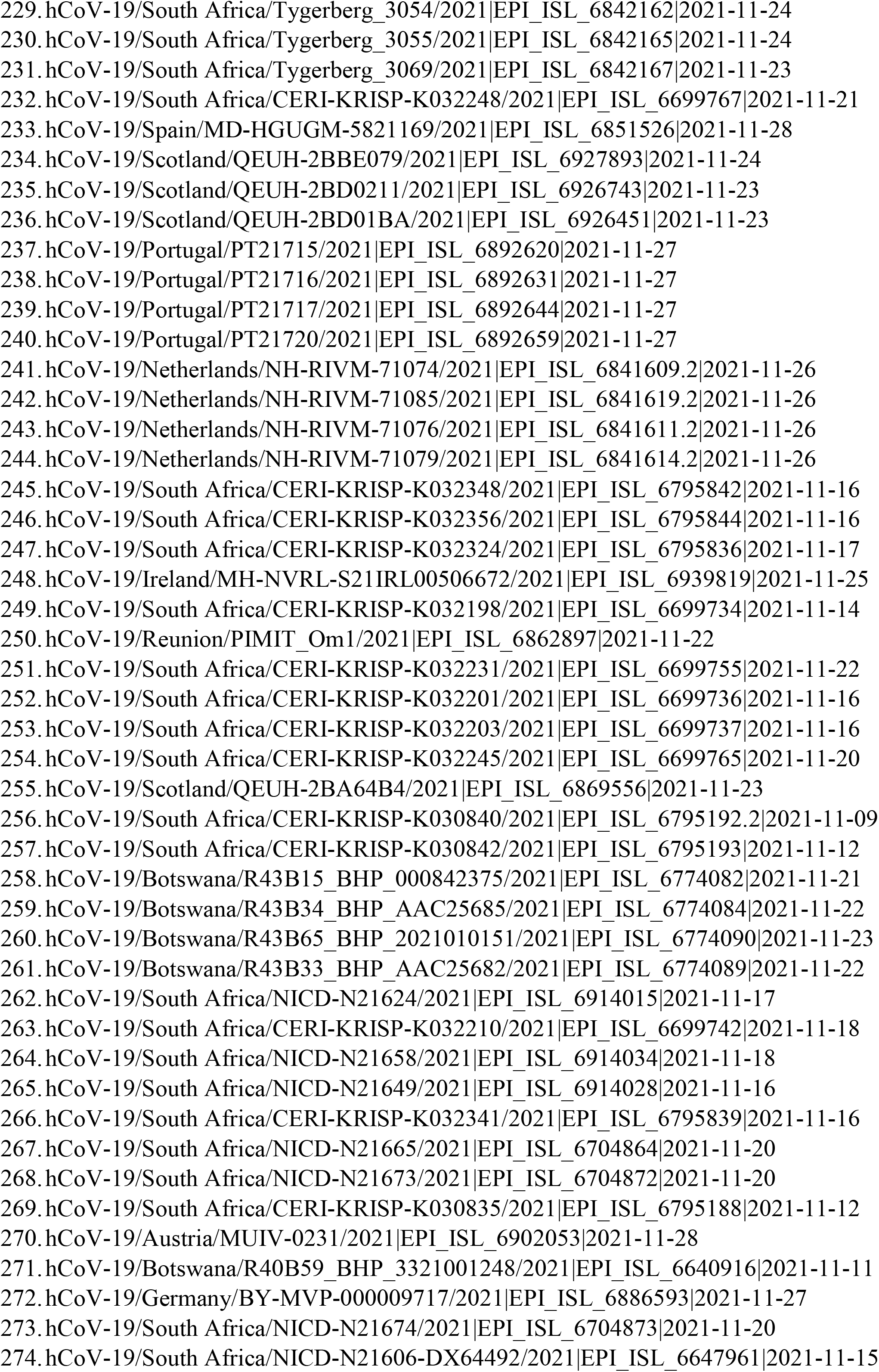
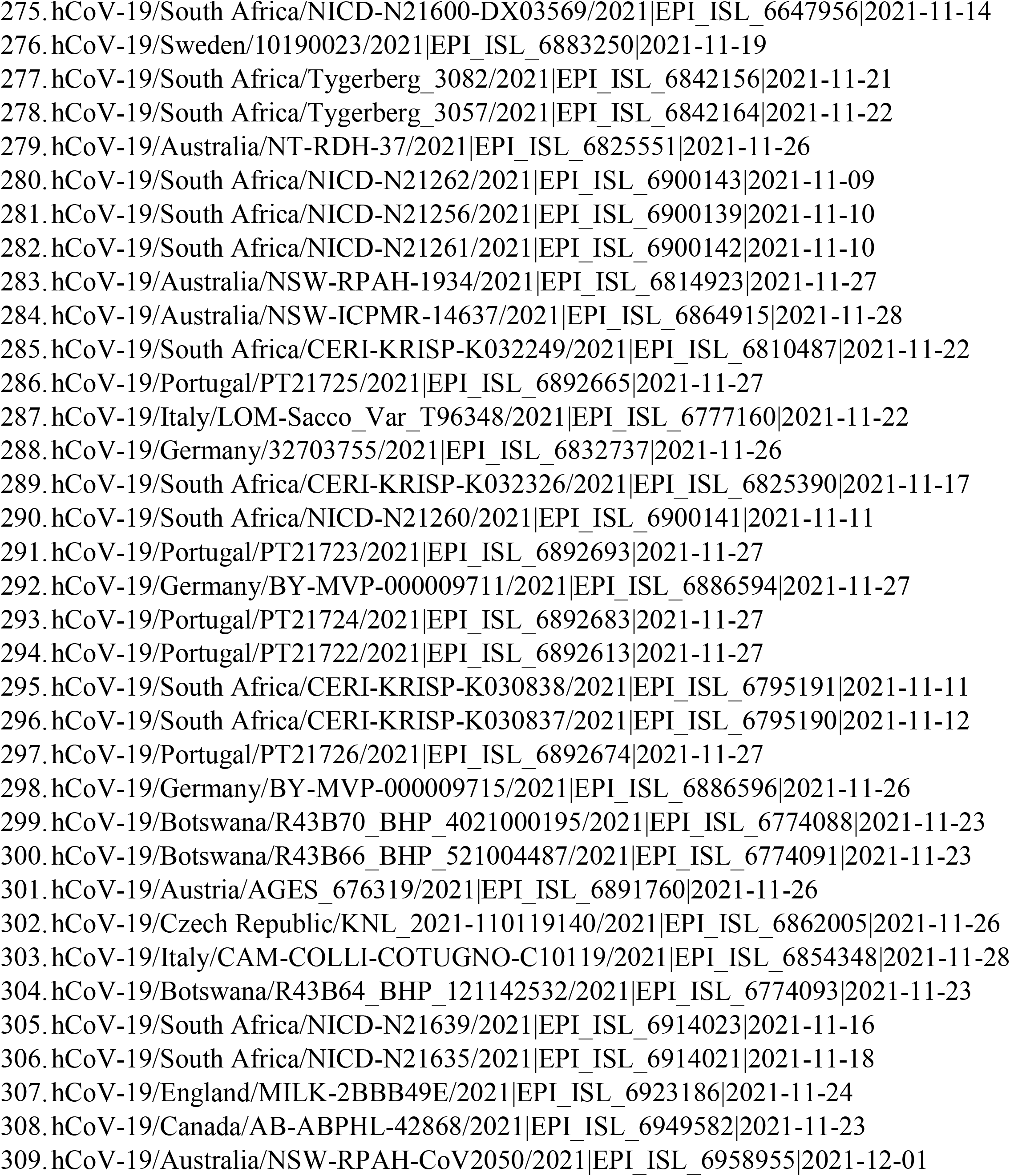
Name of the Omicron variants download from GISAID

**Supplementary Table 2:**
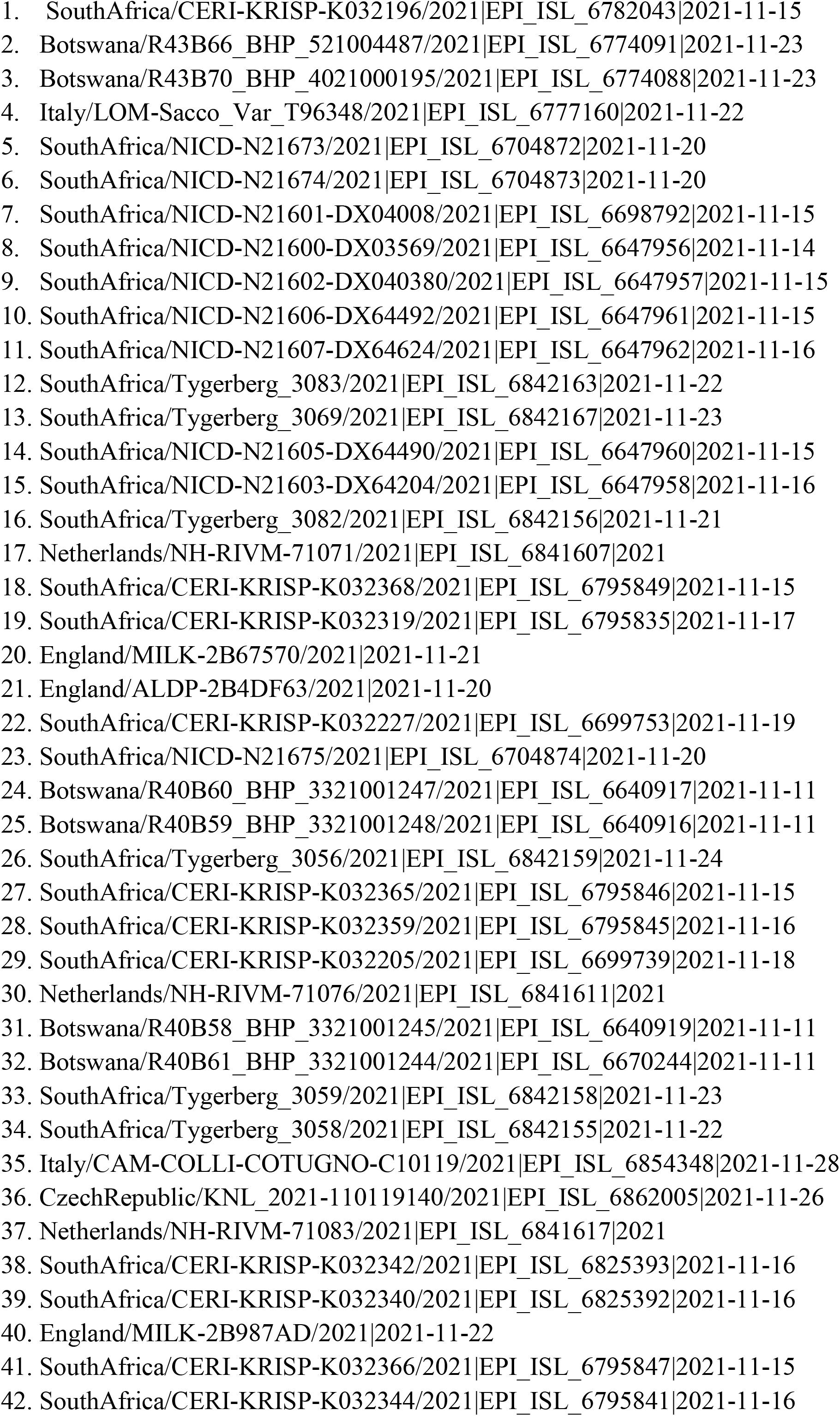
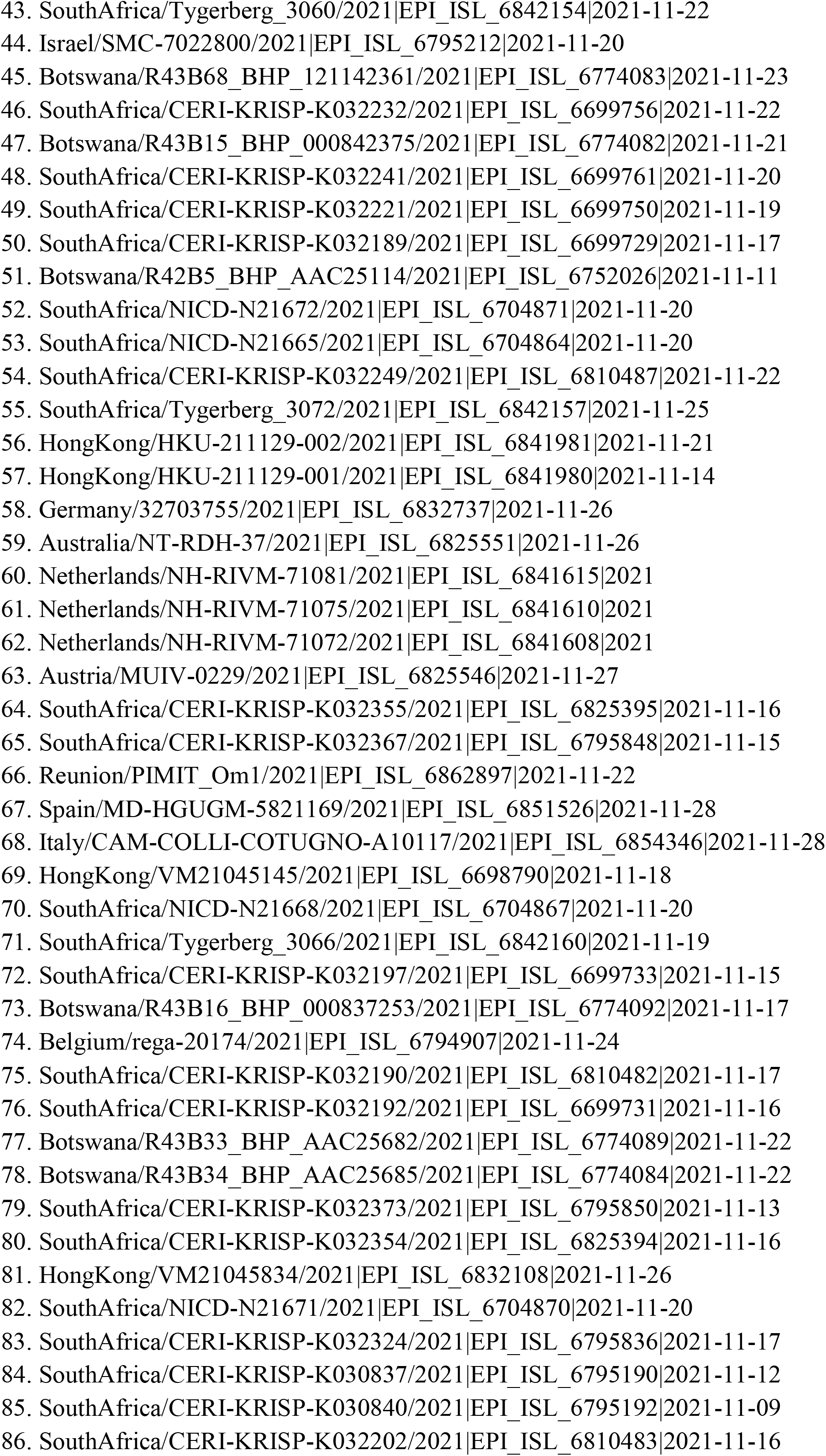
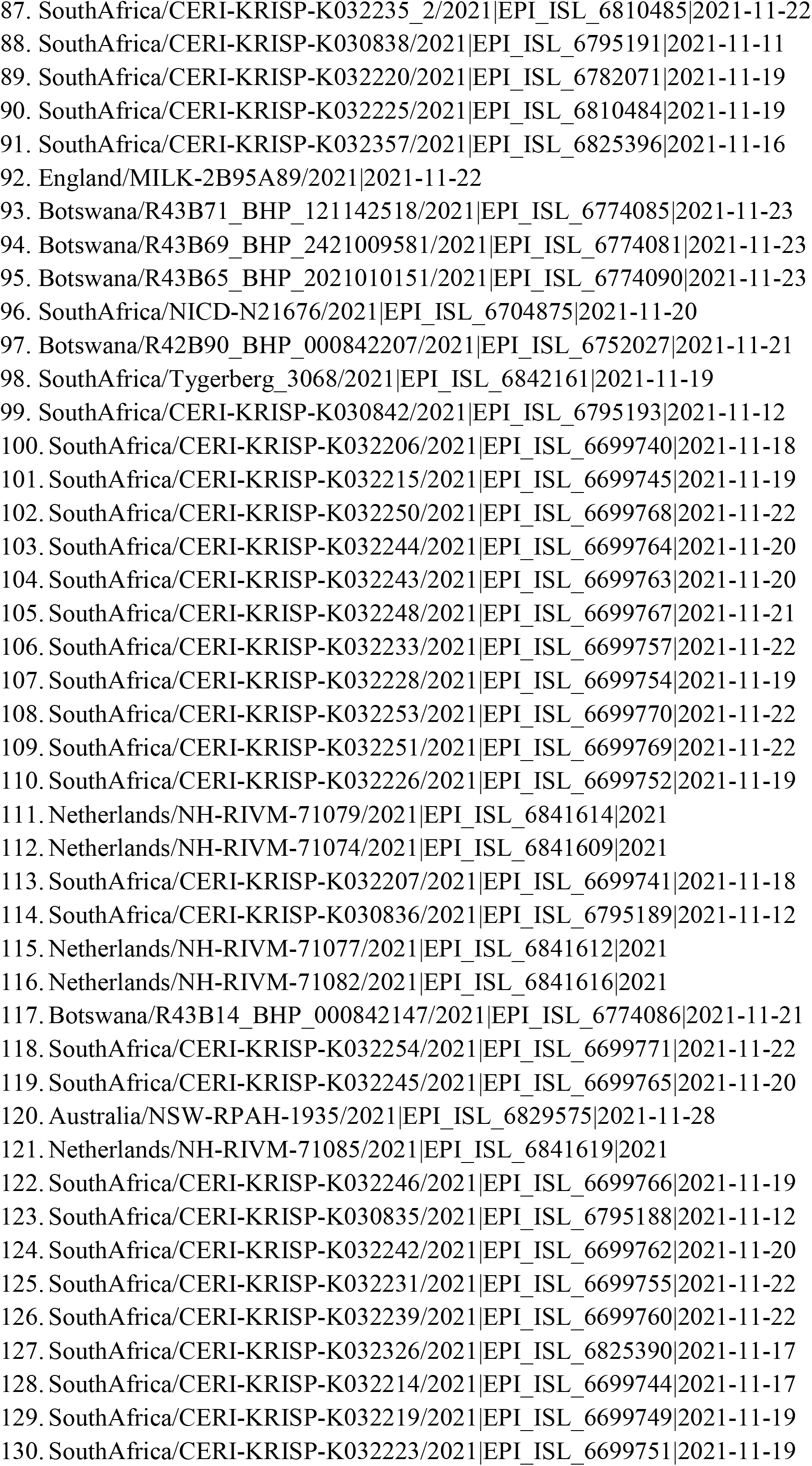
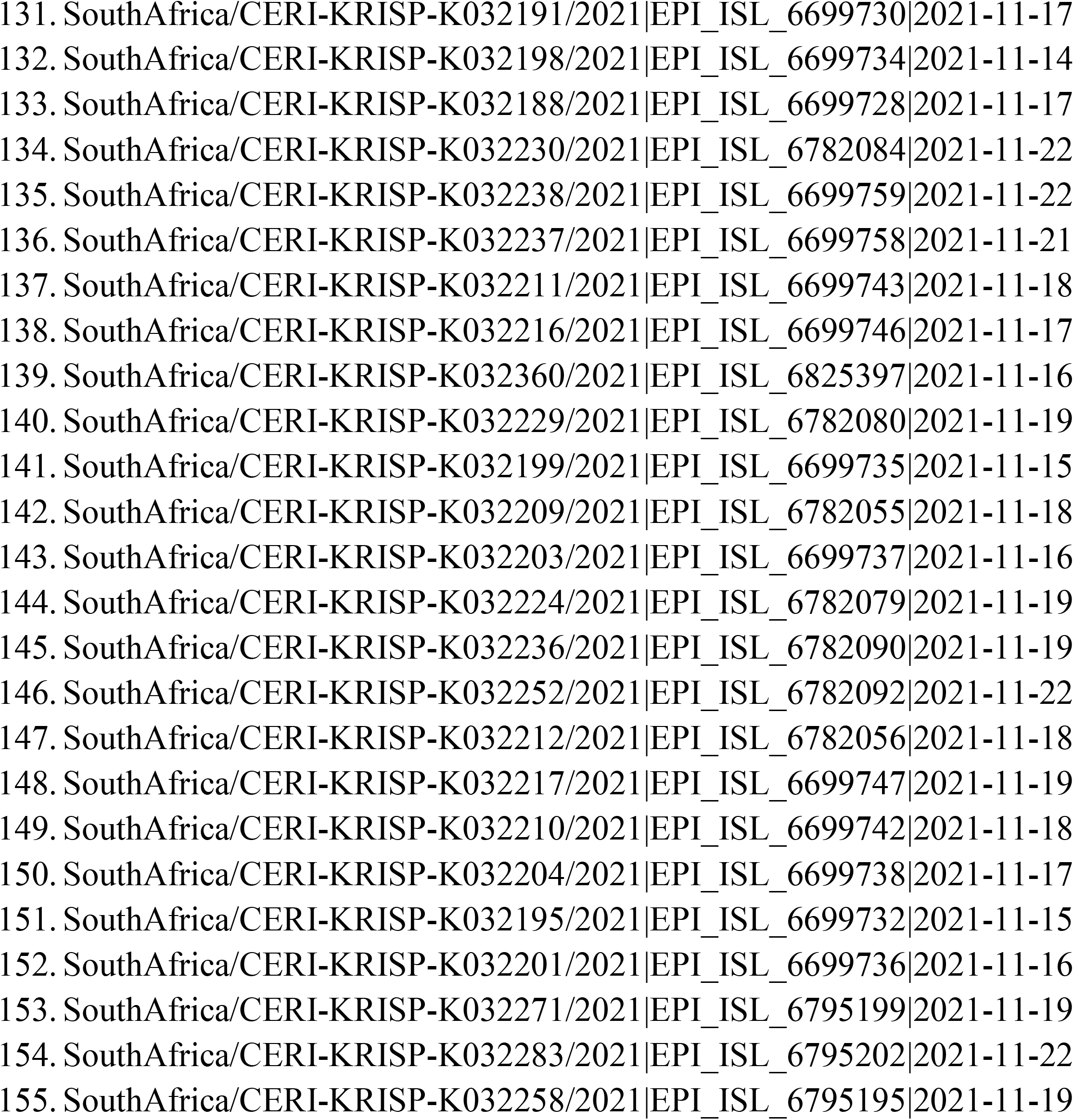
Name of Omicron variants used for the construction of the phylogenetic tree

## References

1. Update on Omicron. World Health Organization (WHO). https://www.who.int/news/item/28-11-2021-update-on-omicron

2. Classification of Omicron (B.1.1.529): SARS-CoV-2 Variant of Concern. World Health Organization (WHO). https://www.who.int/news/item/26-11-2021-classification-of-omicron-(b.1.1.529)-sars-cov-2-variant-of-concern

3. Walls AC, Park YJ, Tortorici MA, Wall A, McGuire AT, Veesler D. Structure, function, and antigenicity of the SARS-CoV-2 spike glycoprotein. Cell. 2020 Apr 16;181(2):281–92.

4. Hoffmann M, Kleine-Weber H, Schroeder S, Krüger N, Herrler T, Erichsen S, Schiergens TS, Herrler G, Wu NH, Nitsche A, Müller MA. SARS-CoV-2 cell entry depends on ACE2 and TMPRSS2 and is blocked by a clinically proven protease inhibitor. cell. 2020 Apr 16;181(2):271–80.

5. Letko M, Marzi A, Munster V. Functional assessment of cell entry and receptor usage for SARS-CoV-2 and other lineage B betacoronaviruses. Nature microbiology. 2020 Apr;5(4):562–9.

6. Zhou P, Yang XL, Wang XG, Hu B, Zhang L, Zhang W, Si HR, Zhu Y, Li B, Huang CL, Chen HD. & Shi, ZL (2020). A pneumonia outbreak associated with a new coronavirus of probable bat origin. Nature.;579(7798):270–3.

7. Wang S, Qiu Z, Hou Y, Deng X, Xu W, Zheng T, Wu P, Xie S, Bian W, Zhang C, Sun Z. AXL is a candidate receptor for SARS-CoV-2 that promotes infection of pulmonary and bronchial epithelial cells. Cell research. 2021 Feb;31(2):126–40.

8. McCallum M, De Marco A, Lempp FA, Tortorici MA, Pinto D, Walls AC, Beltramello M, Chen A, Liu Z, Zatta F, Zepeda S. N-terminal domain antigenic mapping reveals a site of vulnerability for SARS-CoV-2. Cell. 2021 Apr 29;184(9):2332–47.

9. Lempp FA, Soriaga LB, Montiel-Ruiz M, Benigni F, Noack J, Park YJ, Bianchi S, Walls AC, Bowen JE, Zhou J, Kaiser H. Lectins enhance SARS-CoV-2 infection and influence neutralizing antibodies. Nature. 2021 Oct;598(7880):342–7.

10. Walls AC, Tortorici MA, Snijder J, Xiong X, Bosch BJ, Rey FA, Veesler D. Tectonic conformational changes of a coronavirus spike glycoprotein promote membrane fusion. Proceedings of the National Academy of Sciences. 2017 Oct 17;114(42):11157–62.

11. Piccoli L, Park YJ, Tortorici MA, Czudnochowski N, Walls AC, Beltramello M, Silacci-Fregni C, Pinto D, Rosen LE, Bowen JE, Acton OJ. Mapping neutralizing and immunodominant sites on the SARS-CoV-2 spike receptor-binding domain by structure-guided high-resolution serology. Cell. 2020 Nov 12;183(4):1024–42.

12. Starr TN, Czudnochowski N, Liu Z, Zatta F, Park YJ, Addetia A, Pinto D, Beltramello M, Hernandez P, Greaney AJ, Marzi R. SARS-CoV-2 RBD antibodies that maximize breadth and resistance to escape. Nature. 2021 Sep;597(7874):97–102.

13. Tortorici MA, Czudnochowski N, Starr TN, Marzi R, Walls AC, Zatta F, Bowen JE, Jaconi S, Di Iulio J, Wang Z, De Marco A. Broad sarbecovirus neutralization by a human monoclonal antibody. Nature. 2021 Sep;597(7874):103–8.

14. Pinto D, Park YJ, Beltramello M, Walls AC, Tortorici MA, Bianchi S, Jaconi S, Culap K, Zatta F, De Marco A, Peter A. Cross-neutralization of SARS-CoV-2 by a human monoclonal SARS-CoV antibody. Nature. 2020 Jul;583(7815):290–5.

15. Harvey WT, Carabelli AM, Jackson B, Gupta RK, Thomson EC, Harrison EM, Ludden C, Reeve R, Rambaut A, Peacock SJ, Robertson DL. SARS-CoV-2 variants, spike mutations and immune escape. Nature Reviews Microbiology. 2021 Jul;19(7):409–24.

16. McCallum M, Walls AC, Sprouse KR, Bowen JE, Rosen LE, Dang HV, De Marco A, Franko N, Tilles SW, Logue J, Miranda MC. Molecular basis of immune evasion by the delta and kappa SARS-CoV-2 variants. Science. 2021 Aug 11:eabl8506.

17. Shu Y, McCauley J. GISAID: Global initiative on sharing all influenza data–from vision to reality. Eurosurveillance 2017;22(13):30494.

18. Sarkar R, Mitra S, Chandra P, Saha P, Banerjee A, Dutta S, Chawla-Sarkar M. Comprehensive analysis of genomic diversity of SARS-CoV-2 in different geographic regions of India: an endeavour to classify Indian SARS-CoV-2 strains on the basis of co-existing mutations. Archives of Virology 2021;166(3):801–12.

19. Turakhia Y, Thornlow B, Hinrichs AS, De Maio N, Gozashti L, Lanfear R, Haussler D, Corbett-Detig R. Ultrafast Sample placement on Existing tRees (UShER) enables real-time phylogenetics for the SARS-CoV-2 pandemic. Nature Genetics 2021;53(6):809–16.

20. Greaney AJ, Starr TN, Gilchuk P, Zost SJ, Binshtein E, Loes AN, Hilton SK, Huddleston J, Eguia R, Crawford KH, Dingens AS. Complete mapping of mutations to the SARS-CoV-2 spike receptor-binding domain that escape antibody recognition. Cell host & microbe. 2021 Jan 13;29(1):44–57.

21. Arora P, Pöhlmann S, Hoffmann M. Mutation D614G increases SARS-CoV-2 transmission. Signal Transduction and Targeted Therapy. 2021 Mar 1;6(1):1–2.

22. Plante JA, Liu Y, Liu J, Xia H, Johnson BA, Lokugamage KG, Zhang X, Muruato AE, Zou J, Fontes-Garfias CR, Mirchandani D. Spike mutation D614G alters SARS-CoV-2 fitness. Nature. 2021 Apr;592(7852):116–21.

23. Liu L, Wang P, Nair MS, Yu J, Rapp M, Wang Q, Luo Y, Chan JF, Sahi V, Figueroa A, Guo XV. Potent neutralizing antibodies against multiple epitopes on SARS-CoV-2 spike. Nature. 2020 Aug;584(7821):450–6.

24. Barnes CO, Jette CA, Abernathy ME, Dam KM, Esswein SR, Gristick HB, Malyutin AG, Sharaf NG, Huey-Tubman KE, Lee YE, Robbiani DF. SARS-CoV-2 neutralizing antibody structures inform therapeutic strategies. Nature. 2020 Dec;588(7839):682–7.

25. Chi X, Yan R, Zhang J, Zhang G, Zhang Y, Hao M, Zhang Z, Fan P, Dong Y, Yang Y, Chen Z. A neutralizing human antibody binds to the N-terminal domain of the Spike protein of SARS-CoV-2. Science. 2020 Aug 7;369(6504):650–5.

26. Watanabe Y, Berndsen ZT, Raghwani J, Seabright GE, Allen JD, Pybus OG, McLellan JS, Wilson IA, Bowden TA, Ward AB, Crispin M. Vulnerabilities in coronavirus glycan shields despite extensive glycosylation. Nature communications. 2020 May 27;11(1):1–0.

27. Tegally H, Wilkinson E, Giovanetti M, Iranzadeh A, Fonseca V, Giandhari J, Doolabh D, Pillay S, San EJ, Msomi N, Mlisana K. Detection of a SARS-CoV-2 variant of concern in South Africa. Nature. 2021 Apr;592(7854):438–43.

28. Wang Z, Schmidt F, Weisblum Y, Muecksch F, Barnes CO, Finkin S, Schaefer-Babajew D, Cipolla M, Gaebler C, Lieberman JA, Oliveira TY. mRNA vaccine-elicited antibodies to SARS-CoV-2 and circulating variants. Nature. 2021 Apr;592(7855):616–22.

29. Starr TN, Greaney AJ, Addetia A, Hannon WW, Choudhary MC, Dingens AS, Li JZ, Bloom JD. Prospective mapping of viral mutations that escape antibodies used to treat COVID-19. Science. 2021 Feb 19;371(6531):850–4.

30. Weisblum Y, Schmidt F, Zhang F. Fuggite da anticorpineutralizanti da varianti di proteine spike SARS-CoV-2. eLife. 2020;9:e61312.

31. Andreano E, Piccini G, Licastro D, Casalino L, Johnson NV, Paciello I, Dal Monego S, Pantano E, Manganaro N, Manenti A, Manna R. SARS-CoV-2 escape from a highly neutralizing COVID-19 convalescent plasma. Proceedings of the National Academy of Sciences. 2021 Sep 7;118(36).

32. Liu Z, VanBlargan LA, Bloyet LM, Rothlauf PW, Chen RE, Stumpf S, Zhao H, Errico JM, Theel ES, Liebeskind MJ, Alford B. Identification of SARS-CoV-2 spike mutations that attenuate monoclonal and serum antibody neutralization.Cell host & microbe. 2021 Mar 10;29(3):477–88.

33. Walls AC, Miranda MC, Pham MN, Schaefer A, Greaney A, Arunachalam PS, Navarro MJ, Tortorici MA, Rogers K, O’Connor MA, Shireff L. Elicitation of broadly protective sarbecovirus immunity by receptor-binding domain nanoparticle vaccines. bioRxiv. 2021 Jan 1.

34. Tada T, Zhou H, Dcosta BM, Samanovic MI, Mulligan MJ, Landau NR. The Spike Proteins of SARS-CoV-2 B. 1.617 and B. 1.618 Variants Identified in India Provide Partial Resistance to Vaccine-elicited and Therapeutic Monoclonal Antibodies. BioRxiv. 2021 Jan 1.

35. Ferreira IA, Kemp SA, Datir R, Saito A, Meng B, Rakshit P, Takaori-Kondo A, Kosugi Y, Uriu K, Kimura I, Shirakawa K. SARS-CoV-2 B. 1.617 Mutations L452R and E484Q Are Not Synergistic for Antibody Evasion. The Journal of infectious diseases. 2021 Sep 15;224(6):989–94.

36. Li Q, Nie J, Wu J, Zhang L, Ding R, Wang H, Zhang Y, Li T, Liu S, Zhang M, Zhao C. SARS-CoV-2 501Y. V2 variants lack higher infectivity but do have immune escape. Cell. 2021 Apr 29;184(9):2362–71.

37. Cerutti G, Guo Y, Zhou T, Gorman J, Lee M, Rapp M, Reddem ER, Yu J, Bahna F, Bimela J, Huang Y. Potent SARS-CoV-2 neutralizing antibodies directed against spike N-terminal domain target a single supersite. Cell Host & Microbe. 2021 May 12;29(5):819–33.

38. Suryadevara N, Shrihari S, Gilchuk P, VanBlargan LA, Binshtein E, Zost SJ, Nargi RS, Sutton RE, Winkler ES, Chen EC, Fouch ME. Neutralizing and protective human monoclonal antibodies recognizing the N-terminal domain of the SARS-CoV-2 spike protein. Cell. 2021 Apr 29;184(9):2316–31.

39. Starr TN, Greaney AJ, Hilton SK, Ellis D, Crawford KH, Dingens AS, Navarro MJ, Bowen JE, Tortorici MA, Walls AC, King NP. Deep mutational scanning of SARS-CoV-2 receptor binding domain reveals constraints on folding and ACE2 binding. Cell. 2020 Sep 3;182(5):1295–310.

40. Liu Y, Liu J, Johnson BA, Xia H, Ku Z, Schindewolf C, Widen SG, An Z, Weaver SC, Menachery VD, Xie X. Delta spike P681R mutation enhances SARS-CoV-2 fitness over Alpha variant. BioRxiv. 2021 Aug 13.

41. Xia S, Liu M, Wang C, Xu W, Lan Q, Feng S, Qi F, Bao L, Du L, Liu S, Qin C. Inhibition of SARS-CoV-2 (previously 2019-nCoV) infection by a highly potent pancoronavirus fusion inhibitor targeting its spike protein that harbors a high capacity to mediate membrane fusion. Cell research. 2020 Apr;30(4):343–55.

